## Supplementary Material for "Virtual epilepsy patient cohort: generation and evaluation"

### 1 Supplementary Material

#### 1.1 BIDS structure of the virtual epileptic cohort

The BIDS structure used for the VEC dataset is illustrated in ***Figure S1***.

**VirtualEpilepticCohort**

```
|
|--- sub-001                                       # subject id
|     |--- ses-01                                  # each session corresponds to a simulation type
|     |     |--- ieeg                              # simulated SEEG in BrainVision format
|     |          |                                 # each run corresponds to one seizure
|     |          |--- sub-001_ses-01_task-simulatedseizure_acq-VEPhypothesis_run-01_ieeg.eeg
|     |          |--- sub-001_ses-01_task-simulatedseizure_acq-VEPhypothesis_run-01_ieeg.vhdr
|     |          |--- sub-001_ses-01_task-simulatedseizure_acq-VEPhypothesis_run-01_ieeg.vmrk
|     |          |--- sub-001_ses-01_task-simulatedseizure_channels.tsv
|     |
|     |--- ses-02
|     |     |--- ...
|     |
|     |--- sub-001_coordsystem.json                # description on coordinate system
|     |--- sub-001_electrodes.tsv                  # SEEG electrode names and coordinates
|
|--- sub-002
|     |--- ...
|
|--- derivatives                                   # BIDS derivatives format
|     |--- tvb                                     # data simulated using TVB
|           |--- sub-001
|           |     |--- ses-01
|           |     |     |--- VEPhypothesis         # simulation with VEP hypothesis as EZ
|           |     |     |     |--- parameters      # model and simulator parameters in TVB
|           |     |     |     |     |--- sub-001_epileptor_parameters_run-01.tsv
|           |     |     |     |     |---  sub-001_simulator_parameters_run-01.tsv
|           |     |     |     |--- img             #images of the simulated timeseries
|           |     |     |     |     |---  sub-001_simulated_sensor_timeseries_AC_run-01.png
|           |     |     |     |     |---  sub-001_simulated_sensor_timeseries_run-01.png
|           |     |     |     |     |---  sub-001_simulated_source_timeseries_run-01.png
|           |     |     |     |                    # simulated timeseries on the source level
|           |     |     |     |---  sub-001_simulated_source_timeseries_run-01.tsv
|           |     |     |
|           |     |     |--- clinicalhypothesis    # simulation with clinical hypothesis as EZ
|           |     |          |--- parameters
|           |     |          |--- img
|           |     |          |--- ...
|           |     |
|           |     |--- ses-02
|           |     |     |--- ...
|           |     |
|           |     |--- struct
|           |           |
|           |           |--- sub-001_connectome.zip    # structural connectivity
|           |           |--- sub-001_gain.tsv          # gain matrix
|           |           |--- img                       # images of the SC and gain matrix
|           |                |--- sub-001_connectome.png
|           |                |--- sub-001_gain.png
|           |                |--- sub-001_sources_sensors.png
|           |
|           | --- sub-02
|                 | --- ...
|
|--- vep_atlas.tsv                                 # mapping node labels to brain region names
|--- participants.tsv                              # participats id list
|--- dataset_description.json                      # basic dataset metadata (namet, etc.)
|--- README                                        # description of dataset content and usage
```

**Figure S1.** BIDS-IEEG structure of the virtual epileptic cohort. The structure contains the simulated SEEG spontaneous seizures, stimulated seizures and interictal spikes in the main folder of each patient (named sub-001, sub-002, etc.). The parameters used to obtain the simulated time series are detailed in the derivatives folder for each of the EZ hypothesis (either VEP hypothesis or clinical hypothesis).

#### 1.2 Example of two different seizure types for the same patient

Two different seizure types from patient 8 of the virtual epileptic cohort are shown in ***Figure S2***. In these cases, the EZ network is the same, but the propagation network (PZ) is different.

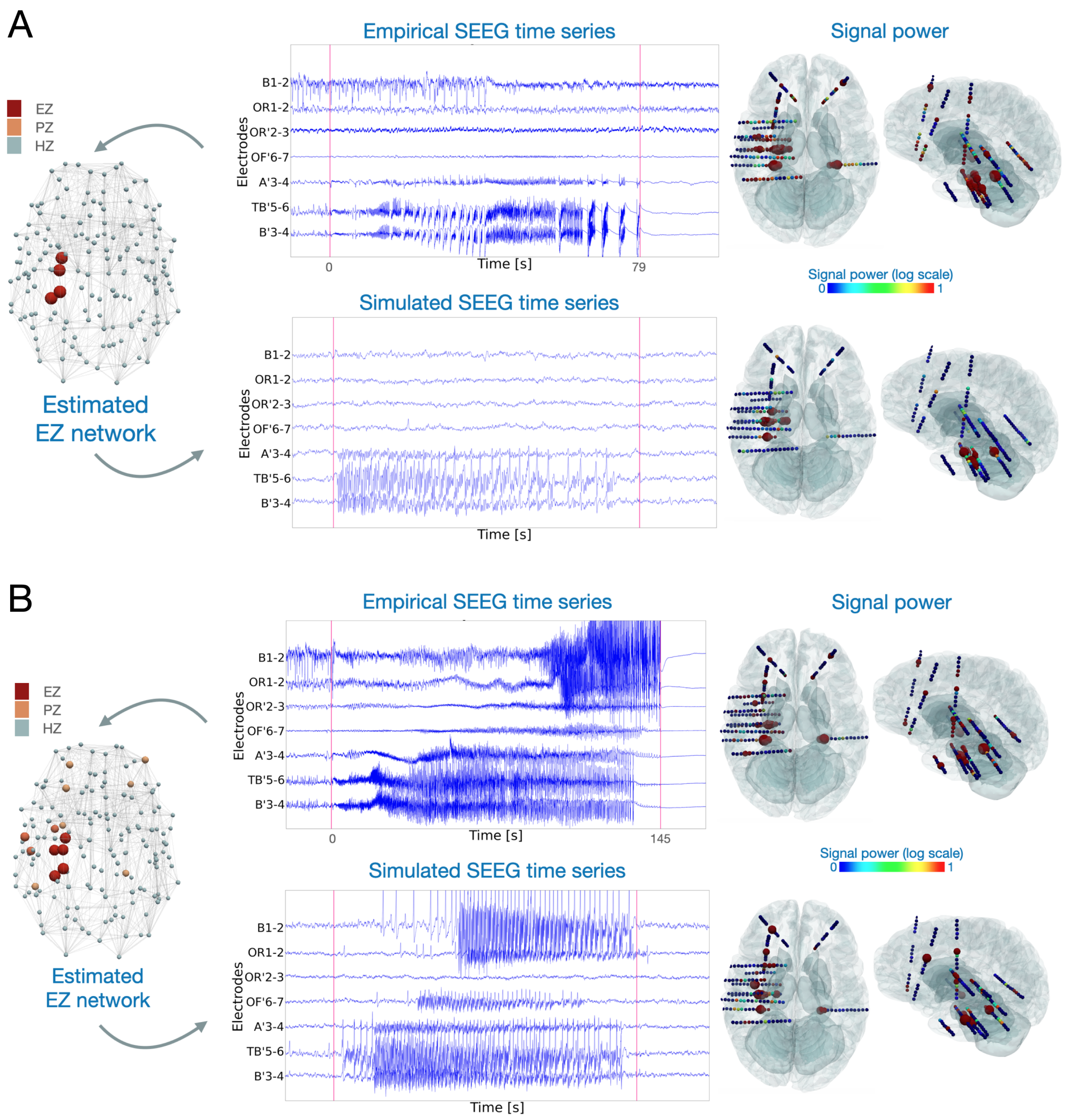


**Figure S2.** Two different seizure types from the same patient. A) Focal seizure occurring in left temporal lobe. B) Propagated seizure with onset in left temporal lobe and propagation to contra lateral hemisphere. For both cases, simulated and empirical SEEG seizures are shown. Red vertical lines indicate seizure onset and seizure offset. Signal power for all SEEG channels is shown in 3D in axial and saggital plane.

#### 1.3 Epileptor-stimulation model

Time series of the main variables in the Epileptor-stimulation model are shown in ***Figure S3*** with 1 Hz stimulation frequency and two different stimulation amplitudes. A higher stimulation amplitude causes a higher increase for the accumulation variable $m$, which can cross the seizure threshold and push the system from the normal state to the seizure state. When the stimulation stops, the variable $m$ slowly returns to its baseline value.

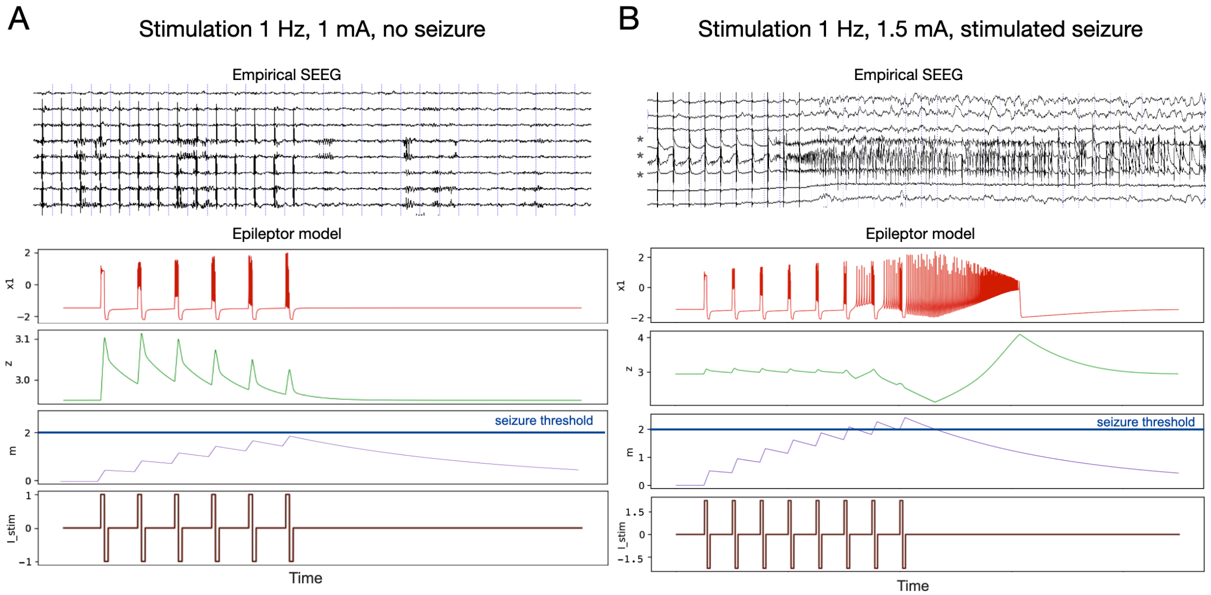


**Figure S3.** Example time series for Epileptor-stimulation model. Above, empirical SEEG recording with stimulation artefact present. Below, simulated time series using the Epileptor-stimulation model. Variables of the model $x_1$, $z$, $m$ and $I_{stim}$ are shown. A) Example with 1 Hz, 1 mA stimulation and no seizure being induced. B) Example with 1 Hz, 1.5 mA stimulation and a seizure is induced. Asterisks indicate the time series containing seizure activity.

#### 1.4 Changing stimulation parameters

An example from patient 10 of the cohort is shown in ***Figure S4*** where different stimulation amplitudes are tested in-silico and compared against the empirical data. Stimulation is applied in electrodes TB'1-TB'2, located in proximity to the left hippocampus region. For all simulations, the EZ hypothesis is estimated from the VEP pipeline and consists of the following regions: Left-Hippocampus-anterior, Left-Hippocampus-posterior, Left-Amygdala. The accumulation hypothesis embedded in the model is such that the effect of the stimulation on the model depends on the seizure threshold. If the critical threshold for seizure onset is reached, the model is kicked to the seizure state (***Figure S4***C) otherwise the model stays in its normal state (***Figure S4***B). Higher stimulation amplitudes will always cross this threshold and destabilise the system to the seizure state (***Figure S4***D).

For the same patient, an example with different stimulation locations is shown in ***Figure S5***. The stimulation location is chosen by randomly selecting a pair of electrodes within a certain radius from the empirical stimulation location. As the stimulus is applied increasingly further away from the empirical stimulation location, the seizure dynamics progressively change from the empirical post-stimulation response.

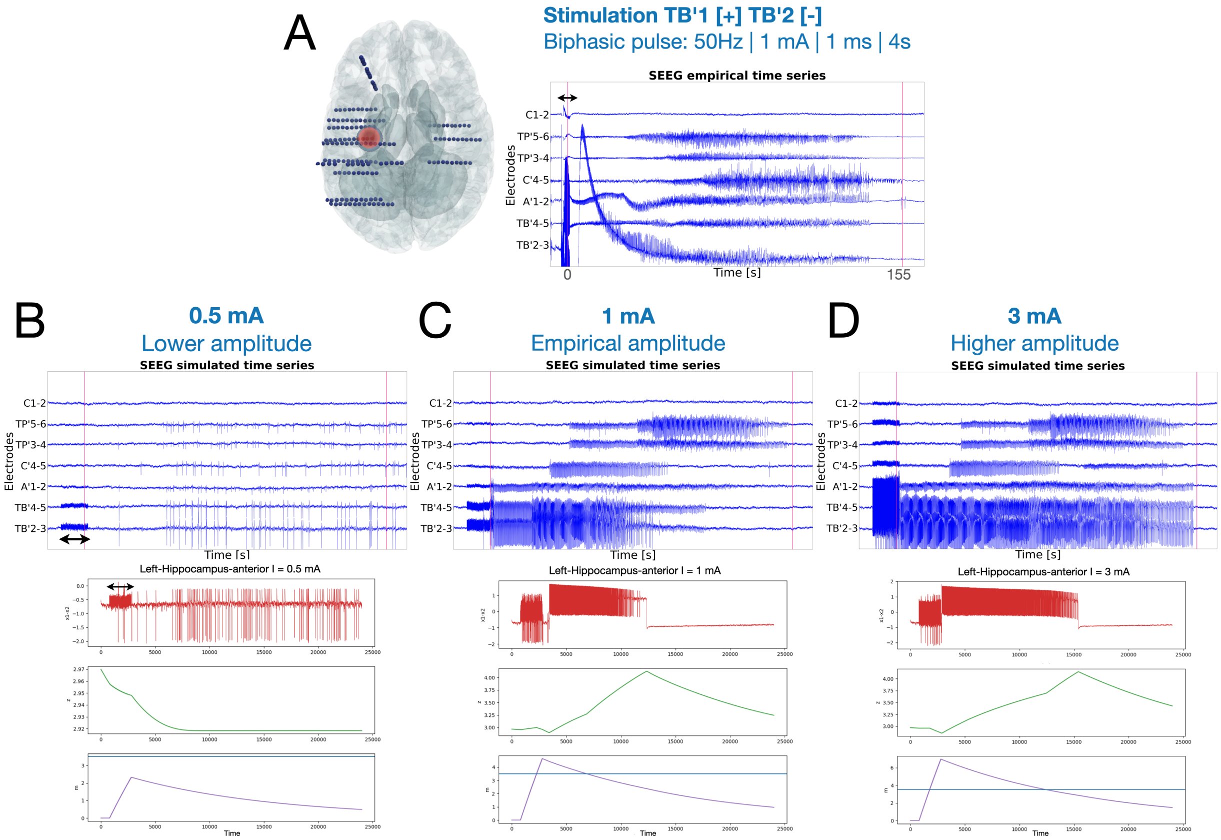


**Figure S4.** Simulation examples with different stimulation amplitude from virtualized patient 10. Double arrow indicates stimulation period. Seven channels are plotted in bipolar montage (out of 116 total bipolar channels). Vertical red lines indicate seizure onset and seizure offset. For all plots B), C) and D), upper plots show simulated time series at the SEEG level with stimulation applied at different amplitudes. Lower plots show variables $x_1 - x_2$ in red, $z$ in green and $m$ in purple evolving for the same simulation for the region left hippocampus anterior. A) Empirical SEEG recording plot of a stimulation-induced seizure. Stimulation was applied at amplitude 1 mA using channels TB'1 [+] and TB'2[-], at frequency 50 Hz, pulse width 1 ms and duration 4 s. Reconstructed SEEG electrodes are shown on the left and stimulation location is plotted in red. B) Upper plot, synthetic SEEG time series of simulated brain activity with stimulation applied at 0.5 mA amplitude. All other stimulation parameters are identical to the empirical parameters. Here, a seizure is not induced after the stimulation is applied. Lower plot, the same simulated activity for the left hippocampus anterior, showing the variable $m$ did not cross the seizure threshold, defined at $3.5$ and corresponding variables staying in the normal state. C) Synthetic SEEG time series of the stimulation-induced seizure at 1 mA amplitude. Here, the same stimulation parameters as the ones applied empirically were used. Following the stimulation, a seizure is induced in the left hippocampus anterior, propagating later on to connected brain structures. Lower plot showing the variable $m$ crossed the seizure threshold and the system is kicked to the seizure state. D) Synthetic SEEG time series plot of simulated brain activity with stimulation applied at 3 mA amplitude. Following the stimulation, a seizure is induced in the left hippocampus anterior and propagating later on to connected brain structures. Lower plot showing the variable $m$ crossed the seizure threshold and the system is kicked to the seizure state.

#### 1.5 Data features for comparing simulated and empirical SEEG

To compare simulated and empirical SEEG, we extracted data features from the SEEG time series. The approach was similar for both the simulated and the empirical cases, and was based on the envelope function, from which the binarized SEEG data were derived (***Figure S6***).

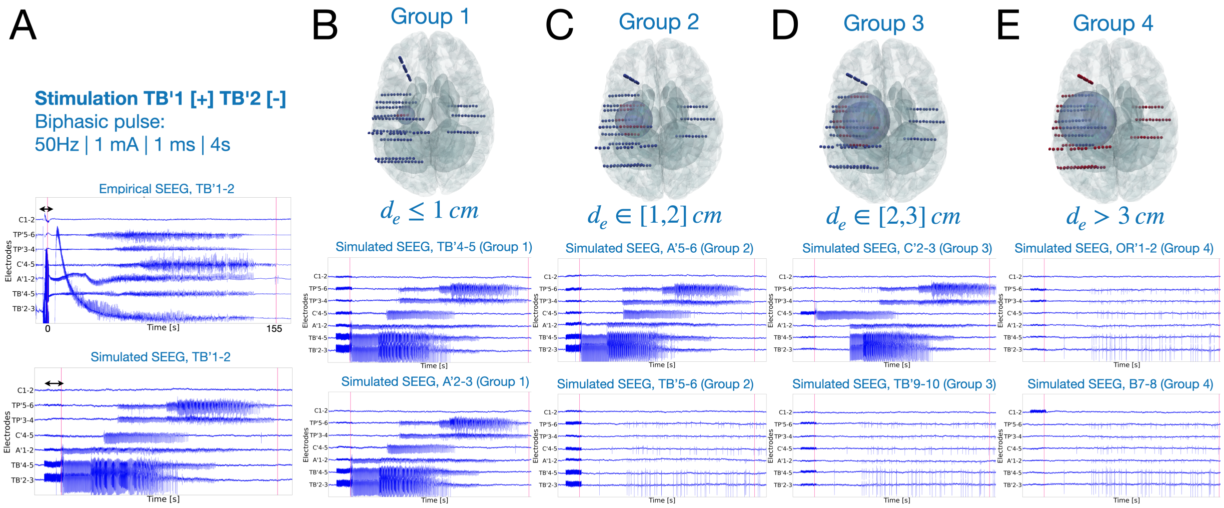


**Figure S5.** Simulation examples with different stimulation location from virtualized patient 10. Double arrow indicates stimulation period. Seven channels are plotted in bipolar montage (out of 116 total bipolar channels). Vertical red lines indicate seizure onset and seizure offset. A) Upper plot, empirical SEEG recording of a stimulation-induced seizure. Stimulation was applied at amplitude 1 mA using channels TB’1 [+] and TB’2[-], at frequency 50 Hz, pulse width 1 ms and duration 4 s. Lower plot, corresponding simulated time series of a stimulation-induced seizure. B) Simulated time series of stimulation applied by electrodes located within 1 cm distance from the empirical stimulation location (TB’1-2). C) Simulated time series of stimulation applied by electrodes located between 1 and 2 cm distance from the empirical stimulation location. D) Simulated time series of stimulation applied by electrodes located between 2 and 3 cm distance from the empirical stimulation location. E) Simulated time series of stimulation applied by electrodes located more than 3 cm away from the empirical stimulation location.

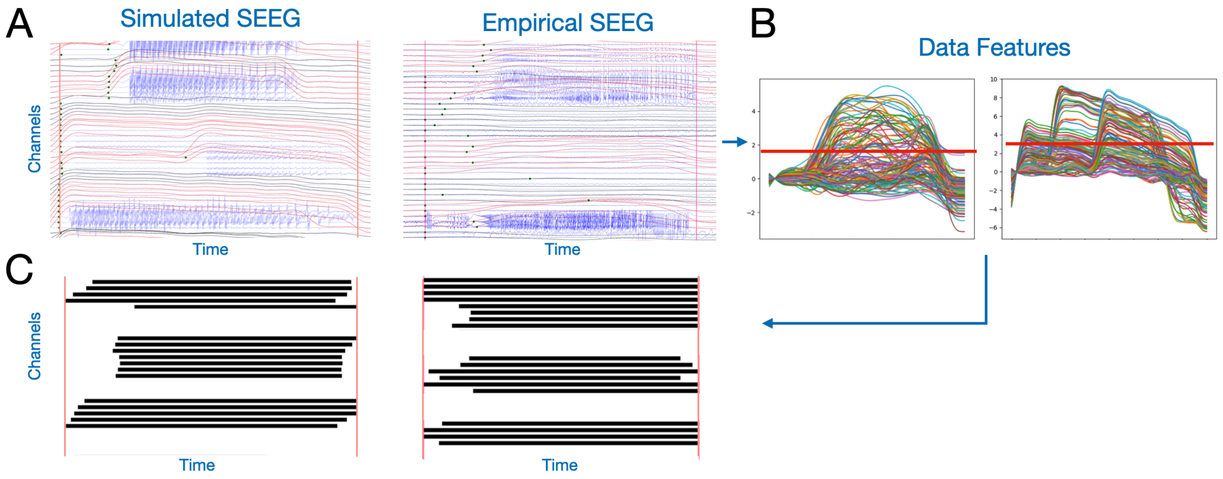


**Figure S6.** Comparing simulated and empirical SEEG: spontaneous seizure example. A) Timeseries plot of a few SEEG channels (right: simulated SEEG time series, left: empirical SEEG time series). Overlayed in red and black are the envelope data features for each SEEG channel, indicating seizure and non seizure channels respectively. Green points indicate estimated seizure onset times. B) Envelope data features overlayed for all SEEG channels (left: simulated, right:empirical). Horizontal red lines indicate chosen threshold to categorize each channel as either seizure (above threshold) or non-seizure channel (below threshold). C) Binary plot of the same SEEG channels, where black indicates seizure activity and white indicates no seizure activity.

#### 1.6 Cohort statistics based on the clinical hypothesis

We used the clinical hypothesis for the EZ ground truth, as an alternative to the VEP hypothesis. We ran the same simulations on the same personalised virtual brain models and computed the similarity metrics as shown in ***Figure S7***. We did not observe a significant difference between the two hypotheses in the computed metrics.

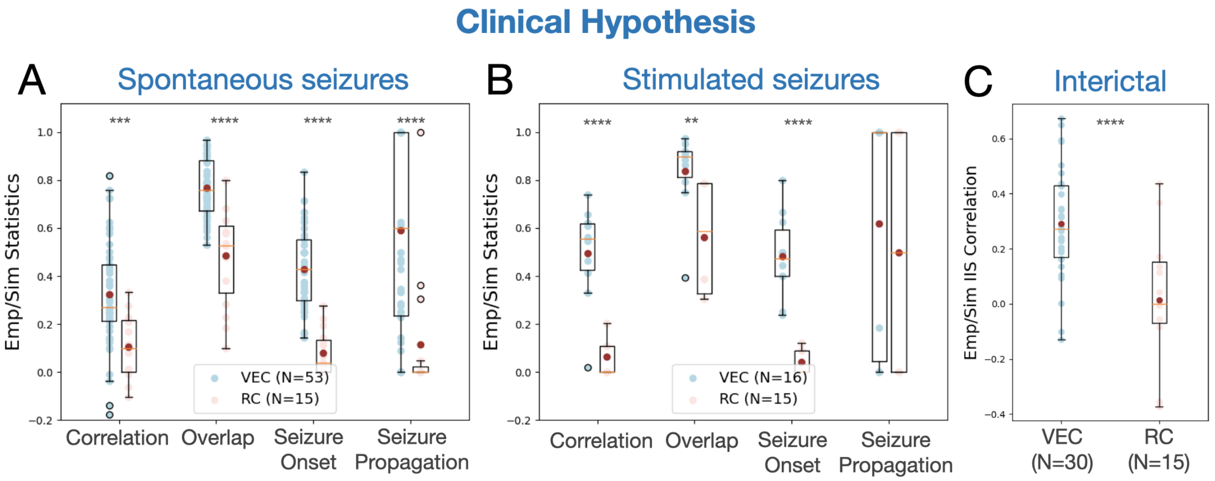


**Figure S7.** Comparison among spontaneous simulated SEEG signals with empirical recordings for the virtual epileptic cohort (VEC, in blue) and the randomized cohort (RC, in red). The clinical hypothesis was used to inform the excitability parameters in the model. A) Boxplot of four main metrics comparing spontaneous seizures against synthetic seizures. Red dots indicate mean values. B) Boxplot of four main metrics comparing synthetic against empirical stimulation-induced seizures. C) Boxplot of interictalspike (IIS) count correlation metric. $^{****}p-value < 0.0001$, $^{***}p-value < 0.001$, $^{**}p-value < 0.01$; permutation test

---

#### 1.7 Complete set of metrics used to compare simulated and empirical seizures

To compare the simulated SEEG time series against the empirical SEEG, we tested sixteen metrics in total (***Figure S8*** and ***Figure S9***). The metrics are explained in the following subsections, following the same order as in the related figures. In the main article, we selected the following metrics: Binary, Jaccard Seizure Onset, Jaccard Seizure Propagation and Correlation.

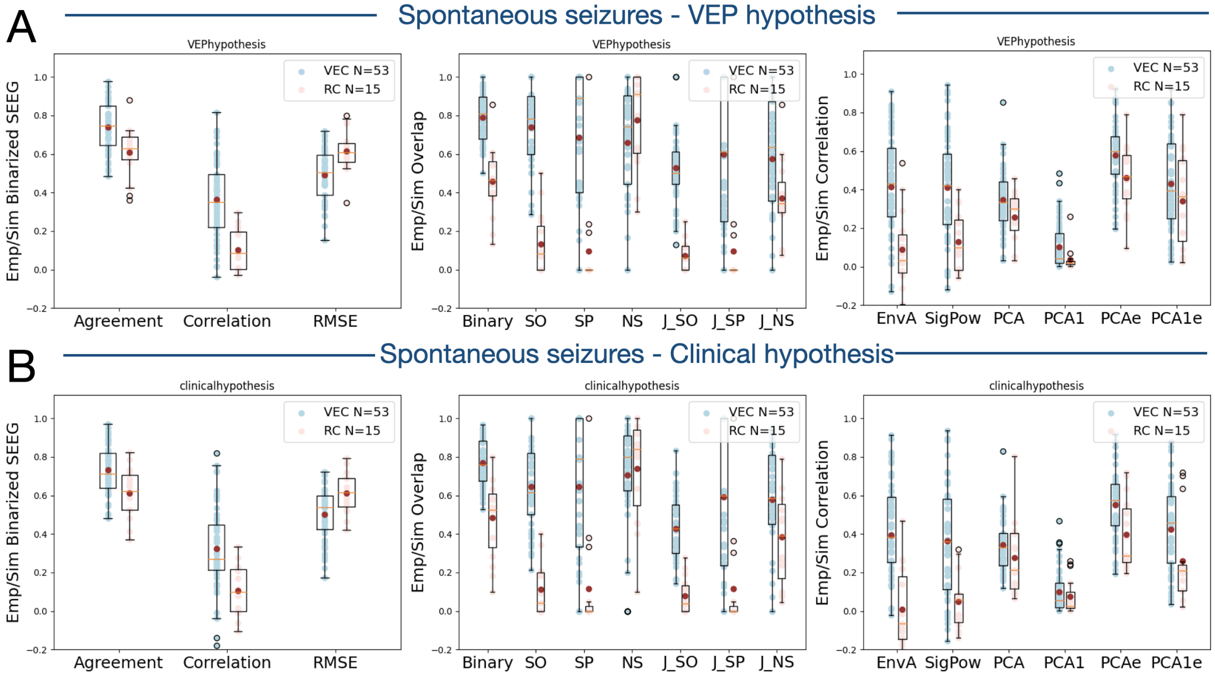


**Figure S8.** Overall statistics for the VEC dataset as compared to the control dataset for spontaneous seizures. In blue, mean and standard deviation for the VEC cohort. In red, mean and standard deviation for the randomized cohort.

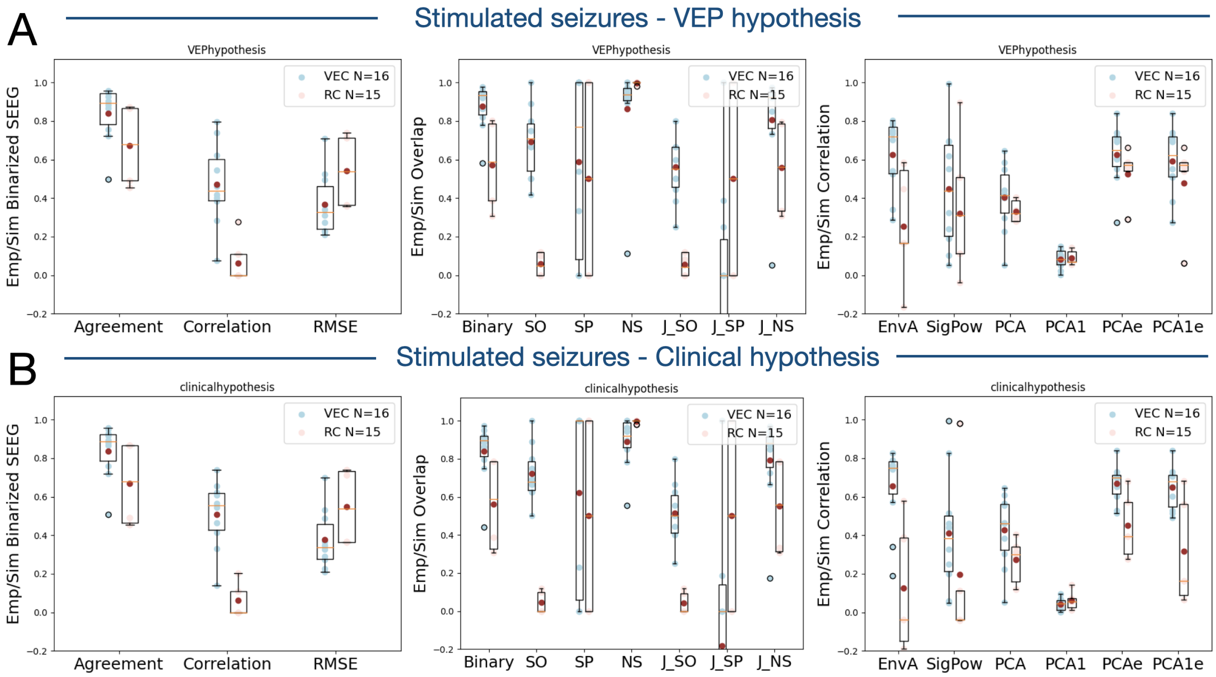


**Figure S9.** Overall statistics for the VEC dataset as compared to the control dataset for stimulated seizures. In blue, mean and standard deviation for the VEC cohort. In red, mean and standard deviation for the randomized cohort.

##### 1.7.1 Overlap metrics

The overlap metric compares, for any given category, the ratio of common SEEG channels between the empirical and simulated sets, divided by the total number SEEG channels in the empirical set. For any given category, it varies between 0 and 1, where 0 indicates no common channels, and 1 indicates that all channels of the empirical set are present in the simulated set.

For both empirical and synthetic SEEG data, we labelled all SEEG channels (in bipolar montage) as either seizure channel ($E_S$, $S_S$ for empirical and simulated case respectively) or no seizure channel ($E_{NS}$, $S_{NS}$ for the empirical and simulated case resp.). A channel is labelled as seizure channel if the envelope data feature crosses the defined threshold, otherwise it is labelled as no seizure channel (as illustrated in ***Figure S6***B). The overlap metric used to compare the synthetic and empirical seizure channels is the following:

$$Binary = \frac{|E_S \cap S_S|}{|E_S|} \tag{1}$$

which computes the ratio between the number of common seizure channels between empirical and simulated SEEG, divided by the total number of empirical seizure channels.

In addition, for both empirical and synthetic SEEG data, we labelled all seizure channels as either seizure onset channel ($E_{SO}$, $S_{SO}$ for empirical and simulated case resp.) or seizure propagation channel ($E_{SP}$, $S_{SP}$ for empirical and simulated case resp.). A seizure channel is labelled as seizure onset channel, if the estimated seizure start time in that channel belongs to the first few seconds of the whole seizure (one-fifth of the total seizure window), otherwise the channel is labelled as seizure propagation channel . The overlap metric used to compare synthetic and empirical data for each category (seizure onset, seizure propagation and no seizure) is as follows:

$$SO = \frac{|E_{SO} \cap S_{SO}|}{|E_{SO}|} \quad SP = \frac{|E_{SP} \cap S_{SP}|}{|E_{SP}|} \quad NS = \frac{|E_{NS} \cap S_{NS}|}{|E_{NS}|} \tag{2}$$

The Jaccard similarity coefficient compares the similarity between to sets, by also taking into account the number of non-common items between the two sets (by computing the union between the two sets). It varies between 0 and 1, where 0 indicates the sets have no elements in common, and 1 indicates the sets are identical.

The Jaccard similarity coefficient applied to each category (seizure onset, seizure propagation and no seizure) is as follows:

$$SO_{Jaccard} = \frac{|E_{SO} \cap S_{SO}|}{|E_{SO} \cup E_{SO}|} \quad SP_{Jaccard} = \frac{|E_{SP} \cap S_{SP}|}{|E_{SP} \cup E_{SP}|} \quad NS_{Jaccard} = \frac{|E_{NS} \cap S_{NS}|}{|E_{NS} \cup E_{NS}|} \tag{3}$$

where $E_{SO}$, $E_{SP}$ and $E_{NS}$ are the empirical seizure onset, seizure propagation and no seizure channels, respectively. $S_{SO}$, $S_{SP}$ and $S_{NS}$ are the synthetic seizure onset, seizure propagation and no seizure channels, respectively.

##### 1.7.2 Correlation metrics

The sample Pearson correlation coefficient is used for all the metrics of this section, where the paired data $(s_1, e_1), ..., (s_n, e_n)$ consisting of $n$ SEEG channels, is used to compute $r$. The coefficient varies between $-1$ and $1$.

$$r = \frac{\sum_{i=1}^{n}(s_i - \bar{s})(e_i - \bar{e})}{\sqrt{\sum_{i=1}^{n}(s_i - \bar{s})^2}\sqrt{\sum_{i=1}^{n}(e_i - \bar{e})^2}}$$

For both simulated and empirical cases, the envelope peak amplitude is computed for each SEEG channel. Then, the sample Pearson correlation between the two lists of amplitude values is computed, $EnvA$.

For both simulated and empirical cases, the signal variance is computed for each SEEG channel. The sample Pearson correlation between the two lists of variance values is computed.

$$V = \frac{1}{T}\sum_{t=1}^{T}(x_t - \bar{x})^2 \tag{4}$$

For both simulated and empirical cases, PCA is performed on the data. First the SEEG timeseries are standardized for each channel, by substracting the mean and dividing by the standard deviation. Then, PCA is performed on the time series using the same number of components as the number of SEEG channels. Then, the Pearson correlation is computed between the main principal components (i.e. the ones that explain 90% of the variance) of the empirical and simulated data, thus obtaining a correlation matrix. From this matrix, the maximum correlation value is extracted as a comparative value, $PCA$. As a second approach, the Pearson correlation is computed between the two first components of the empirical and simulated data, $PCA1$.

For both simulated and empirical cases, PCA is performed on the envelope data features, using the exact same approach as above, but replacing the SEEG time series by the envelope time series. The metrics computed are $PCA_e$ and $PCA1_e$.

##### 1.7.3 Comparative metrics for 2D binarized SEEG

We performed four main comparative metrics on the binarized images ($E_{bin}$ for the empirical case, $S_{bin}$ for the simulated case) obtained from the SEEG time series (as illustrated in ***Figure S6***C).

First, we performed an overlap metric by measuring the number of identical binary (simulated, empirical) pairs divided by the total number of pixels for one image.

$$agreement = \frac{|E_{bin} \cap S_{bin}|}{|E_{bin}|} \tag{5}$$

Second, we performed a Pearson $correlation$ on the 2D images, by comparing each binary (simulated, empirical) pair of identical row and column.

Third, we computed the mean squared error and the root mean squared error between the two binary images, with total number of $N$ values:

$$mse = \frac{1}{N}\sum_{i=1}^{N}(E_{bin}(i) - S_{bin}(i))^2 \quad rmse = \sqrt{mse} \tag{6}$$

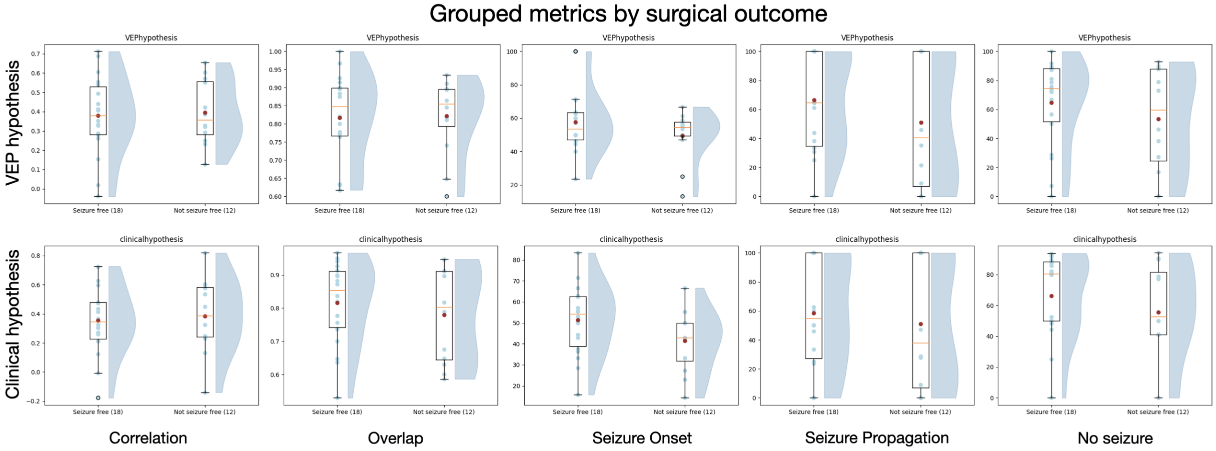


**Figure S10.** Grouped metrics following surgical outcome. Patients were grouped in either the seizure-free group or the not-seizure-free group. First row shows metrics from synthetic data using the VEP hypothesis. The second row uses the clinical hypothesis. In each plot, metrics from seizure-free patients are plotted on the left side, wherease metrics from not-seizure-free patients are plotted on the right side. A boxplot is overlaid over all individual data points and a violin plot from the same data points is shown on the right next to it.

#### 1.8 Grouping metrics by surgical outcome

We grouped the patients of the VEC cohort according to the surgical outcome in two groups: seizure-free and not-seizure-free. In figure S10, we plot the metrics performance for these two groups. These data were pooled following the Engel score of each patient, where patients with Engel score I were assigned the seizure-free group and patients with Engel scores II, III and IV were assigned the not-seizure-free group. This analysis was purely performed for exploratory purposes. Our interpretation follows that since our purpose was to build sythetic data that best match the empirical recordings, we seem to have little to no bias in whether the data came from patients that were seizure-free or not.
